# Daily Healthy Eating Index (HEI-2020) scoring reveals diet quality patterns masked by aggregation

**DOI:** 10.64898/2026.06.08.26355152

**Authors:** Rohan Singh, Marcel Salathé

## Abstract

The Healthy Eating Index (HEI-2020) is conventionally computed by aggregating intake across days before scoring. Digital food logging enables an alternative: scoring each day and averaging daily scores. These methods are not equivalent. The HEI’s density-based structure and component caps cause aggregation to inflate adequacy scores when intake is irregular. Using Food & You data, we show daily HEI correlates more strongly with microbiome diversity, and recommend co-reporting both metrics.

## 1) Introduction

### 1.1 Background and Structure of the Healthy Eating Index

The Healthy Eating Index (HEI) was introduced in 1995 to provide a single indicator of overall diet quality for nutrition monitoring and policy evaluation^1^, with updates released to align with successive editions of the Dietary Guidelines. Major revisions in 2005 and 2010 shifted the index toward a density□based, food pattern approach. HEI□2015 introduced added sugars and saturated fat as moderation components, and HEI□2020 retained the component structure and standards while aligning the name with the 2020-2025 guidance.

The HEI□2020 is a 13-component, 0 to 100 summary measure of overall diet quality that reflects adherence to the Dietary Guidelines for Americans (DGA)^2^. Components are classified as adequacy components, for which higher intake is encouraged, or moderation components, for which lower intake is recommended; for both, higher scores reflect closer alignment with the guidance. Most components are scored on a density basis per 1,000 kcal with proportional scoring between minimum and maximum standards. The fatty acids component is an exception to this density approach and is scored as a ratio of poly- and monounsaturated fatty acids (PUFAs and MUFAs) to saturated fatty acids (SFAs), with higher ratios indicating better diet quality. The total score is the sum of component scores and provides a comparable metric across individuals, menus, or food supplies.

### 1.2 Clinical Utility and Health Associations

HEI is expected to be a predictor of long-term health outcomes across multiple domains. In a recent dose□response meta□analysis of 18 published studies, higher HEI□2015 scores were linked to about 20% lower all□cause mortality, with similar reductions for cardiovascular and cancer mortality and a non□linear dose□response^3^. Meta-analyses that pool HEI with other dietary indices underscoring clinical utility show consistent risk reductions in various health outcomes, including approximately 22% lower all□cause and cardiovascular risk, 16% for cancer, 18% for type 2 diabetes, and 15% for neurodegenerative disease^4^. Furthermore, NHANES US data support this pattern, showing lower HEI-2015 associated with higher stroke prevalence, with significantly stronger protective effects observed in women than men^5^. Beyond cardiometabolic outcomes, HEI also shows better health associations in bone^6^, frailty^7^, and oral health^8^. In microbial ecology, HEI also captures meaningful biology, explaining the greatest variance in gut microbiome alpha and beta diversity among common indices in a twin cohort^9^.

### 1.3 Conventional Scoring Approaches

The index can be applied to 24hr recalls, food records, menus, and FFQs, though the method choice depends on the research question and acceptable error structure. The NCI (National Cancer Institute) provides documentation for multiple approaches, including per□person scoring on multi□day totals and day□specific scoring^10^.

The purpose of HEI is to capture usual diet quality rather than that of a single day, because intake varies from day to day. In standard practice, when multiple days of dietary data are available per subject, the standard recommended approach is to aggregate (sum) intakes and energy across all days for that subject, then compute densities (e.g. grams per 1,000 kcal) and apply the scoring algorithm once to yield a single “usual diet” HEI score. For population-level means (when only one 24-hour recall per subject is available), the population ratio method is preferred because it pools intakes and energy across individuals before computing ratios, reducing bias from day-to-day variability. When replicate intake data exist, NCI suggests bivariate or multivariate usual-intake methods that can refine estimates by modeling and removing within-person variation and accounting for correlation among dietary components and generate regression-calibrated estimates^10^. Under all these approaches, higher total or component HEI scores reflect closer alignment with dietary guidelines - and because HEI is based on density (intake per energy), it captures quality of diet rather than absolute quantity.

However, these methods were originally developed to correct for day-to-day dietary noise in sparse, nonconsecutive recall data, by modeling within-person variation and inter-component correlation to estimate distributions of habitual HEI scores. But when diet is logged continuously for many successive days (e.g. 2–4 weeks), the internal variation is directly observable, reducing the need for heavy modeling. In such settings, applying NCI-style “shrinkage” or simulation may even obscure temporal features that are meaningful (e.g. day-to-day fluctuations). Therefore, using daily HEI estimates or short-window aggregations becomes plausible and may be more biologically informative in long intake-logging cohorts.

### 1.4 Rationale and Objectives

The aggregate□then□score convention simplifies analysis and aligns with historical reliance on instruments that do not resolve daily variability, especially FFQs. Yet two HEI design features can compress information when intake is irregular: pern1,000 kcal normalization couples component densities to daily energy, and component caps or floors create plateaus once standards are met or missed. Adequacy components often exhibit a concave, capped relationship with intake^13^, so scoring a multi□day average of densities can exceed the average of day□level scores when intake oscillates between very low and very high days. Moderation components behave differently because scoring decreases with intake and has different thresholds. These mechanics motivate co□reporting daily HEI alongside the conventional period average now that daily data are readily captured.

In a recent publication^11^, we computed both overall (average quantity) HEI and average daily HEI and found that average daily HEI correlated more strongly with gut microbiome diversity than the overall HEI. This supports the idea that preserving day-level information is biologically meaningful. Building on this background, we examine how HEI□2020’s scoring structure interacts with day□to□day variability, describe scenarios where an overall HEI can overstate sustained diet quality, and discuss implications for study design in nutritional health research that can increasingly leverage daily diet data.

## 2) Why averaging quantities can “overcompensate”

Adequacy components increase linearly with intake until a maximum threshold is reached, after which the score remains capped. Because of this structure, averaging intakes across days and then applying the scoring function can yield higher values than averaging the daily scores, as demonstrated in Figure 1A. Surplus intake on high-consumption days contributes to the multi-day average intake but is discarded when scoring those days individually due to the daily cap. As a result, the score based on averaged intake can exceed the mean of the daily scores. For example, in a two-day scenario where an adequacy component is not consumed at all on day 1 but is consumed at a very high level on day 2, the daily scores would average to 50%. However, if day 2 intake is at least twice the threshold, the two-day average intake reaches the threshold, and the multi-day method assigns the maximum score.

**Figure 1.**
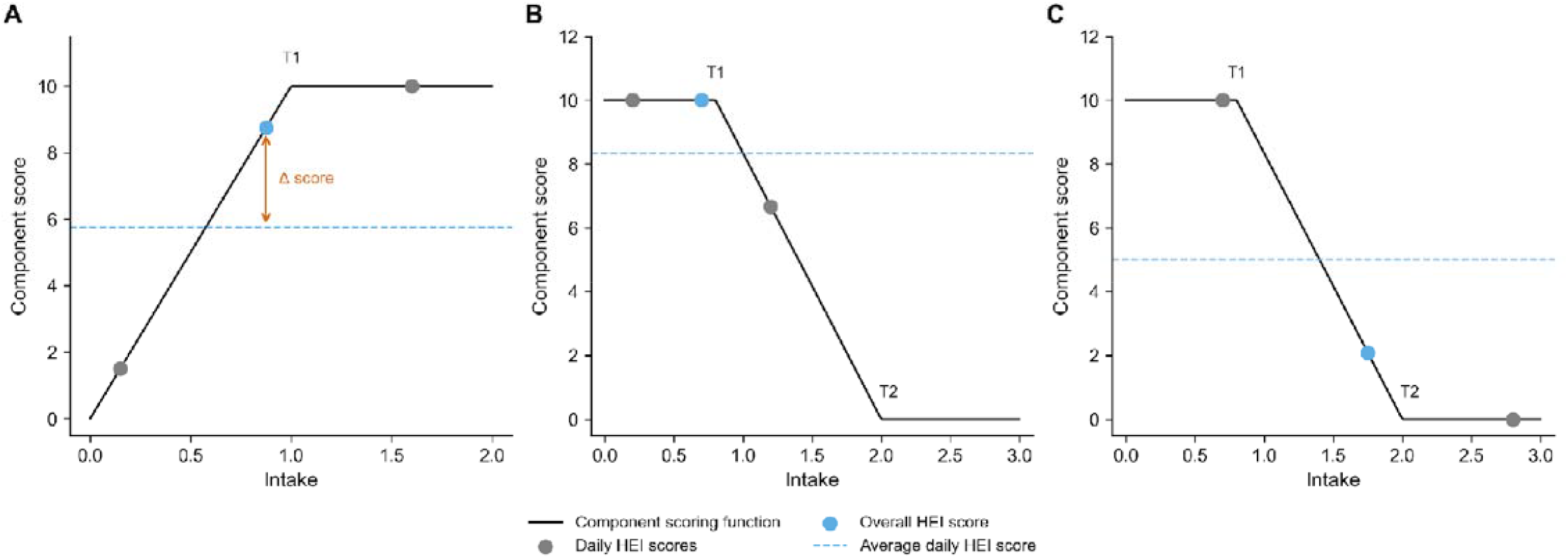
Divergence between aggregate and daily HEI scoring methods driven by thresholded component scoring. Each panel shows a two-day intake scenario with daily HEI component scores (gray dots), the score computed from aggregated intake (blue dot), and the mean of daily scores (dashed blue line). Quantity thresholds used in scoring are explicitly marked (T1, T2). Panel A: Adequacy components increase linearly with intake up to a maximum threshold (T1), beyond which scores are capped. When intake is irregular across days, surplus intake on high-consumption days compensates for deficits on low-consumption days under aggregation, yielding a higher score than the mean of daily scores. Panels B-C: Moderation components receive maximum points below a lower threshold (T1) and decline linearly to zero at an upper threshold (T2). The direction of divergence depends on intake patterns. In (B), a low-intake day offsets a moderately high-intake day, allowing aggregation to remain below T1 and inflate the score relative to the daily average. In (C), a high-intake day exceeds T2, pulling the aggregated intake into the penalized region and reducing the aggregate score relative to the mean daily score.

Moderation components behave differently because their scoring function is flat up to a first threshold (T_1_), then declines linearly to zero at a second threshold (T_2_). Averaging quantities can therefore have two distinct effects. In one case, a day with intake slightly above T_1_ can be offset by a day with very low intake, pulling the average quantity back below T_1_ (as shown in Figure 1B); the multi-day aggregate method would then award a maximum score, while the daily average score would remain below the maximum due to the elevated first day. In the opposite case, a high-intake day can raise the multi-day average sufficiently to enter the declining region between T_1_ and T_2_, reducing the score, whereas the daily average method, with one high day and one low day, may not register this reduction (Figure 1C). Thus, for moderation components, scoring based on averaged intake can yield either higher or lower values than averaging daily scores, depending on the intake pattern.

A separate mechanism can counteract this tendency for adequacy components when energy intake and nutrient density are negatively correlated. Because the original HEI method computes density from total intake divided by total energy, it effectively weights each day’s contribution by that day’s energy. Days with higher energy intake exert greater influence on the final score than days with lower energy intake. In contrast, the mean daily method assigns equal weight to every day regardless of energy. When an individual tends to consume nutrient-dense meals on low-energy days and nutrient-poor meals on high-energy days, the aggregate method downweights the favorable days while amplifying the unfavorable ones. The mean daily method does not impose this weighting and therefore preserves the full contribution of the low-energy, high-density days. If this negative correlation is sufficiently strong, it can overwhelm the ceiling effect described above, resulting in a mean daily score that exceeds the score derived from aggregated intake.

The energy-weighting mechanism also applies to moderation components. Because the aggregate method weights each day’s contribution by its energy intake, the relationship between energy and nutrient density influences the outcome independently of threshold effects. If high-energy days tend to have lower densities of moderation components (favorable), the aggregate method amplifies these days and may yield a higher score than the mean daily method. Conversely, if high-energy days coincide with higher densities of sodium, saturated fat, or added sugars, the aggregate method penalizes this pattern while the mean daily method preserves equal contribution from the more favorable low-energy days. Thus, the direction of the difference depends on the individual’s behavioral patterns, and this energy-weighting effect combines with the threshold-related effects to determine the net discrepancy between the two scoring approaches.

## 3) Results

Figure 2 illustrates the divergence between overall and daily HEI scores across individual components in Food & You, a digital cohort that collected continuous dietary intake data via the mobile food logging app MyFoodRepo in Switzerland from 2018 to 2023^12^.

**Figure 2.**
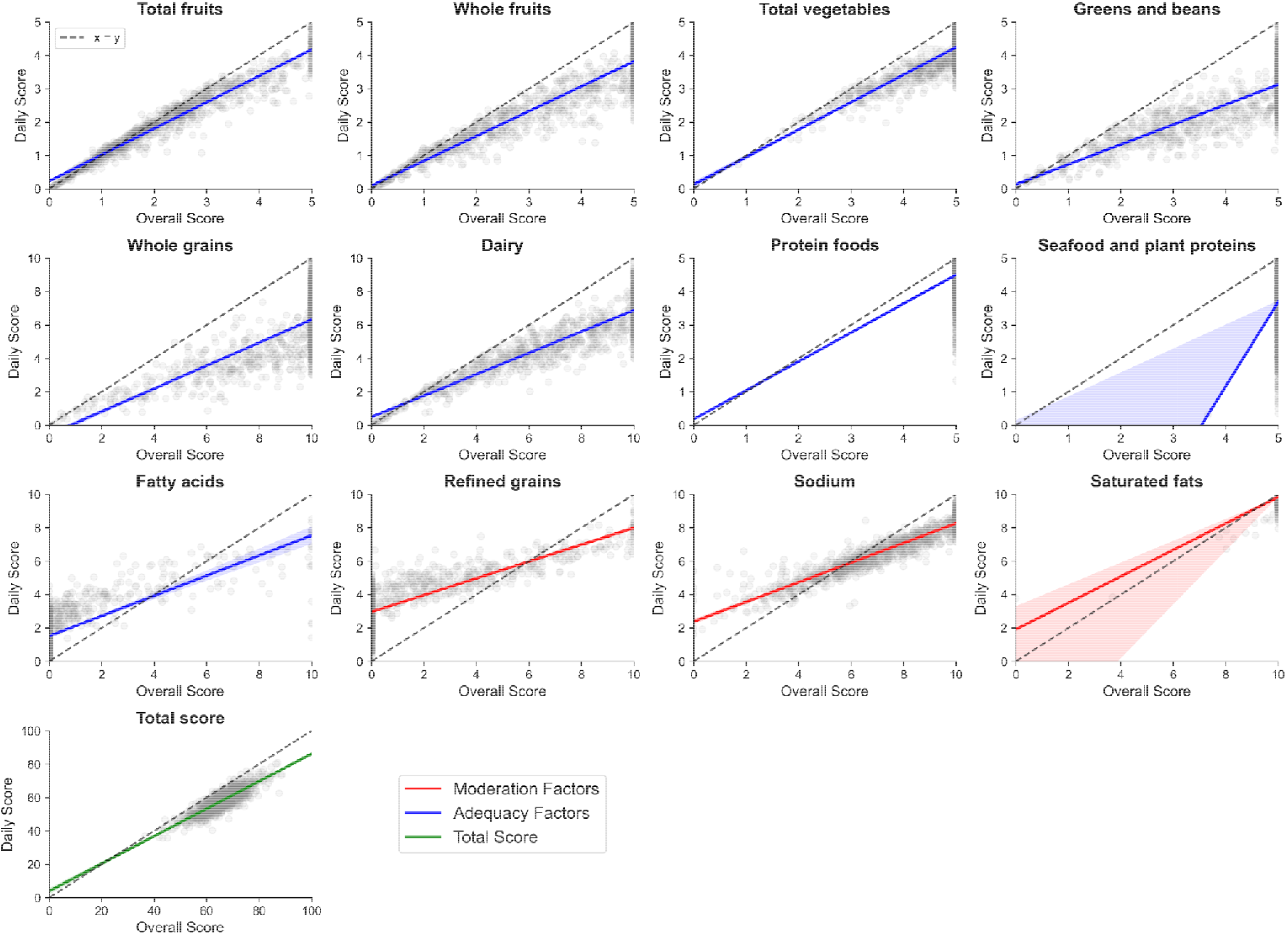
Comparison of daily versus overall Healthy Eating Index (HEI-2020) scores for each component and the total score in the Food & You digital cohort. Each point represents participant’s mean daily score (y-axis) against their score computed after aggregating intakes across all days (x-axis). Blue panels show adequacy components, red panels show moderation components, and green represents the total score. The dashed line indicates the line of equality (y = x), and shaded bands denote 95% confidence intervals around fitted regression lines.

HEI-2020 component and total scores were computed for each participant following Kohl et al.^13^ under two approaches. The overall (aggregate) score was obtained by summing intakes and energy across all of a participant’s logging days, computing densities on these totals, and applying the scoring algorithm once. The mean daily score was obtained by applying the scoring algorithm to each logging day individually and averaging across the participant’s 14- or 28-day tracking period^12^; following the cohort’s dietary handling, days with total energy intake below 1,000 kcal were excluded^12^. Each panel plots the mean of daily HEI scores (y-axis) against the corresponding overall HEI score (x-axis) per individual, with the dashed identity line representing equivalence between the two approaches, a fitted regression line, and its 95% confidence band. Across components, we observe deviation from the equivalence line, highlighting how ceiling effects, threshold-based scoring, and energy-weighting interact to produce non-equivalent estimates under the two approaches in real-world data as explained above.

### 3.1 Adequacy Components: Overestimation From Aggregation

For most adequacy components, including Total Fruits, Whole Fruits, Total Vegetables, Greens and Beans, Whole Grains, Dairy, Protein Foods, and Seafood and Plant Proteins, the fitted relationship lies consistently below the identity line. This indicates that daily HEI scores are, on average, lower than scores derived from aggregated intake. The pattern is strongest at higher overall scores, where aggregation pools intake across days to surpass density thresholds, while daily scoring is constrained by component-specific ceilings. Excess intake on high-consumption days therefore increases the aggregate density but does not yield additional daily points once the maximum score is reached. Nevertheless, dispersion around the fitted line reveals heterogeneity in individual intake patterns. A minority of observations lie above the identity line, suggesting cases where daily averages exceed aggregate scores. These deviations are consistent with negative correlations between energy intake and nutrient density, whereby high-quality intake occurs on lower-energy days. Because the aggregate method weights days by energy intake, such high-density, low-energy days contribute less to the overall score, producing lower aggregate values relative to the unweighted daily mean. The Fatty Acids component, scored as the ratio of poly- and monounsaturated to saturated fatty acids rather than as an intake density, behaves differently: its fitted relationship sits above the identity line at lower scores. Without the per-1,000 kcal ceiling that drives overestimation in the other adequacy components, aggregation does not produce the same systematic gap between daily and aggregate scoring.

### 3.2 Moderation Components: Reversed Bias Pattern

Moderation components exhibit the opposite pattern. For Sodium and Refined Grains, the regression lines sit above the identity line, indicating that daily HEI scores tend to exceed aggregate scores. This reversal arises from the structure of moderation scoring, where intake below a first threshold receives full points and scores decline linearly beyond a second threshold. When intake oscillates around these thresholds, averaging quantities before scoring can push the aggregate density into the penalized region, whereas averaging daily scores preserves the contribution of favorable low-intake days. However, as discussed above, the energy-weighting mechanism can modulate this effect: if high-energy days coincide with lower densities of moderation components, the aggregate method may yield comparable or even higher scores than the daily average. Saturated Fats shows wider dispersion, with the fitted relationship crossing the identity line rather than sitting consistently above it, indicating that the direction of divergence is not uniform across participants and depends on individual intake patterns.

### 3.3 Total Score

At the summary level, the Total Score panel shows the same downward deviation observed in adequacy components, confirming that overall HEI is typically higher when derived from aggregated intake. This reflects the dominance of adequacy components in driving the total score difference, as their ceiling effects outweigh the opposing bias from moderation components.

## 4) Implications for research in the era of daily tracking

Together, these results highlight how HEI-2020’s capped, density-based design interacts with energy variability to inflate period-averaged diet quality. These effects are largest for episodically consumed adequacy foods (e.g., greens, legumes, whole grains) and for participants with high energy amplitude. By contrast, daily HEI captures temporal irregularity—information now observable through digital diet logs—that the aggregate method compresses.

The conventional period-averaged HEI remains well-suited for research questions concerning cumulative dietary exposure and chronic disease risk. However, the two scoring approaches are not interchangeable, and the choice between them has implications for inference. In research domains where physiological systems respond to short-term dietary inputs, day-to-day consistency in diet quality may carry information beyond the period average.

The gut microbiome is an example of such a domain. In our recent analysis of the Food & You cohort, daily HEI correlated more strongly with microbiome alpha diversity than period-averaged HEI^11^, suggesting that sustained regularity in diet quality, not just its average level, may shape the microbial ecosystem. For studies with multi-day intake records, we recommend co-reporting period-averaged and mean daily HEI. This approach preserves comparability with existing literature while retaining temporal information that digital food logging now makes available at scale.

Several practical applications follow. In intervention studies, daily HEI could distinguish treatments that improve average diet quality from those that also stabilize it. Two interventions producing identical period-averaged gains may differ in whether participants maintain quality consistently or oscillate between adherent and non-adherent days. Daily HEI variability metrics could also identify dietary phenotypes invisible to aggregated scoring, such as individuals who compensate for poor weekend eating with high weekday quality versus those who maintain moderate quality throughout; both groups may share similar average scores yet differ in metabolic or microbiome outcomes. In dietary trials, the standard deviation of daily HEI could complement mean adherence as a measure of compliance stability. More broadly, individuals with high day-to-day variability may benefit from different nutritional guidance than those with consistently low or high diet quality, opening avenues for tailored intervention strategies.

There are some limitations. Because the dietary composition databases did not separate added sugars from intrinsic sugars, the HEI-2020 Added Sugars component was approximated using total sugars and is not shown among the components in Figure 2. Continuous daily logging, and thereby the daily HEI used here, is not feasible in all study designs and is best suited to cohorts and interventions with consecutive daily intake records.

## Data Availability

Metadata containing clinical, demographic, and nutritional variables cannot be deposited publicly due to participant privacy and ethical restrictions. Access to this metadata can be requested by contacting the corresponding author, subject to institutional ethical compliance.

